# TREATMENT OUTCOMES AND ASSOCIATED FACTORS AMONG SEVERELY MALNOURISHED UNDER-FIVE CHILDREN ADMITTED TO JINKA GENERAL HOSPITAL, SOUTH ARI ZONE, SOUTH E THIOPIA REGION

**DOI:** 10.1101/2024.11.05.24316770

**Authors:** Turufat Tukura

**Author notes:** Author’s.

## Abstract

**Introduction:** Recurrent drought has contributed to household level food insecurity that resulted in malnutrition in the zone. This contributed to high prevalence of children malnutrition.

At the Jinka General Hospital the treatment outcomes and associated factors were not identified by research where there are high admissions of children with severe acute malnutrition with significant mortality. Therefore, this study was aimed to conduct a retrospective review of records of children admitted with severe acute malnutrition to Jinka General Hospital to assess treatment outcomes and associated factors at stabilization centre

**Objective:** This study was to assess treatment outcomes and associated factors in children with severe acute malnutrition admitted to Jinka General Hospital.

**Method:** A hospital record-based data analysis study was conducted to assess the treatment outcomes and associated factors of severe acute malnutrition among a total of 453 children aged 0 to 6 months and 6–59 months old. Severe acute malnutrition registration book and patient charts was used as a source of data. Data was entered into STATA version 16 for analysis. Ordered logistic regression was used to analyse treatment outcome with successful (cured), death (poor) and unknown (unsatisfactory). Variables at P-value <0.05 were considered as predictors of treatment outcome among children admitted with Severe Acute Malnutrition (SAM).

**Results:** Rehydration Solution for Malnutrition (ResoMal), educational status of mother, exclusive breast feeding, returned defaulter, severe wasting with bilateral Pitting oedema (Marasmus-Kwashiorkor), Infants 0-6 Months of age, and Unconscious were the substantial predictors of death among children admitted with SAM. Whereas therapeutic milk F-100, Gentamycin, sex of the child (male), rural residence, new admission, bilateral Pitting Oedema +++(Kwashiorkor), and some dehydrations were the predictors of recovery among the SAM cases.

**Conclusion:** exclusive breast feeding, and educational status of mother are the prevention of malnutrition. Targeted intervention of cases based on the enter category to stabilization centres is imperative for better outcome. The study also highlighted that the cure rate was below the SPHERE treatment outcome standard and death rate was at alarming rate according to SPHERE standard and national treatment outcome indicator. Thus, the study recommends management of SAM children through adherence with National Guideline for the Management of Acute Malnutrition in Ethiopia and empowering of women and girls to improve their literacy. This will contribute to improve household level food preparation practice and services to prevent malnutrition among children.

## Introduction

Severe malnutrition is the most common cause of morbidity and mortality among children under-five years of age. Many children with SAM die at home without care, but even when health care is provided, mortality rates may still be high. Health care providers, mistakenly use practices that are suitable for most non-malnourished children, but highly dangerous for severely malnourished children resulted in usual loss of lives. Efficacy of SAM management protocol defined as how well the protocol works in ideal and controlled settings which measured by cure rate in clinical trial. If full protocol of SAM management implemented including early identification the recovery rate should be close to 100 percent[1, 2].

Study conducted at therapeutic feeding units of Mekelle city indicated unacceptably high case fatality rate among children admitted with SAM in the first week of admission. Similar study conducted at Gedeo zone, indicated that the survival rate reduces with increasing in length of stay (95.3% first week,90% second week, 85% third week). Similar studies conducted in Ethiopia and other parts of the world mostly reported that the case fatality rate was significantly associated with Diarrhoea, IV antibiotic, blood transfusion, IV infusion, hypoglycaemia, shock, altered pulse rate and altered body temperature, type of medical treatment at hospital [3, 4].

Recovery rate of SAM cases management considering WHO protocol is at Hawassa University Referral hospital is below the standard. Blood transfusion, intravenous fluids and age were associated with poor quality of care at the hospital which doubled by severe anaemia and HIV infection[5].

Report of 2016 Ethiopian Demographic and Health Survey (EDHS) indicated that children under five years of age in Ethiopia were stunted (38%), wasted (10%) and underweight (24%). Likewise; in the south Ethiopia region the magnitude of under nutrition in under-five years of age children were stunting(39%), wasting(6%) and underweight(21.1%)[6].

Malnutrition declines economic growth and prolongs poverty through mortality and morbidity that a direct loss in human capital and productivity for economy. Poor cognitive function and reduced school performance indirectly losses the country’s economy. In total, the economic cost of malnutrition is estimated to range from 2to 3 % of GDP, to as much as 16% in most affected countries. Malnutrition has social and economic implications of 11% of a country’s (Gross Domestic Product) GDP loss annually due to the high prevalence of under nutrition [7].

Total losses associated with under nutrition are estimated at ETB 55.5 billion or US$4.7 billion for the year 2009 which are equivalent to 16.5% of GDP of that year in Ethiopia. Among factors associated with child deaths under -nutrition takes 28% and there were an estimated of 378,591 additional annual cases of child mortality associated with child under-nutrition in the period from 2004 to 2009[8].

Nearly 20 million children are estimated to be severely acutely malnourished worldwide. Most of them live in South Asia and in Sub-Saharan Africa including Ethiopia. Malnourished children are prone to 5 to 10 times higher risk of death compared to well-nourished children[9]. Approximately 2.9 million children accessed treatment in 65 countries1n 2013 only about 17% of children taking medical care despite significant improvement in recent years[9].

According to studies conducted in different parts of the world, children admitted with SAM to hospital setting had showed the recovery rate of 30 to 78 % [2-21]. Despite this effort recent systematic review indicated that the case fatality rate for inpatient treatment of SAM ranges from 3.4 to 3.5 percent even if there is hospital admissions for SAM cases management and updating of management protocols[10].

So far, studies conducted in different parts of Ethiopia mostly focused on Children aged above six months age and did not use SPHERE treatment outcome standard as the reference of comparison of treatment outcome. In addition, all studies were conducted at non-pastoral communities.

## Method and Material

### Study design

A Hospital record based retrospective data analysis was conducted.

### Study population and source population

Source populations was records of SAM patients aged 0-59 months of age admitted to Jinka General Hospital stabilization centre. Study population were records of randomly selected eligible children aged 0-59 months of age with SAM admitted to Jinka General Hospital stabilization center.

### Inclusion criteria

Records of all children with SAM treated at Jinka General Hospital stabilization centre were included into the study.

### Exclusion criteria

Children whose socio-demographic variables and admission criteria were not registered in their SC card and Children with age group of >5 years.

### Variables of study

### Dependent variable (outcome)

Treatment outcomes: Successful (cured), dead (poor), unknown (defaulter, transferred out, and non-responded). To simplify the analysis of the study the treatment outcome was further classified as excellent, poor and unknown.

### Independent variables (predicators)

Age, admission type, medication, clinical form of SAM.

### Operational definitions

**Treatment outcomes (Unsuccessful)**: - discharged dead, defaulter, non-responder, transfer out. **Recovered (cured or successful**): -Child that has reached the discharge criteria for inpatient care.

**Transfer out to OTP**: child that is transferred from inpatient care to OTP.

**Defaulter:** - child has absent for 2 consecutive visits in patient care.

**Death: -** patient that has died while s/he was in the in –patient care.

**Non-responder**: - patient that has not reached the discharge criteria after 40 days in the patient care

**Readmission**: -Patient that has defaulted from a nutritional therapeutic treatment, and he is re-admitted into unit within a period of less than 2 months.

**Kwashiorkor**: Bilateral pitting oedema (nutritional oedema) that is a clinical indicator for SAM.

### Sample size and sampling procedure

For the first specific objective, the sample size is determined using single proportion population formula with the following assumptions: an average cure rate in admitted under-five children with SAM to be **77.8%** [18], 95% level of significance, and 5% as margin of error, then it was calculated as follows:

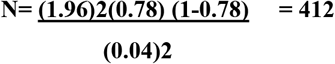

By adding 10% for incomplete data, the final sample size will be 453.

### Sample size for associated factors

| Variable | Assumption | Sample size | Reference |
| --- | --- | --- | --- |
| Age | OR=2, P=74%, Ratio 1:1Power=80%, CI=95% | 452 | Efrem etal,2010 |
| Edema | OR=2, P=89, Ratio=1:1power=80%, CI=95% | 410 | Tefera etal,2014 |

Since the sample size for single population proportion (453) is greater than sample for associated factors (452), the sample size of the study was 453 for the study.

### Sampling procedure

Table of simple random numbers was used to select the required sample size of 453 among the recorded medical records.

### Data collection procedures

Checklist was used for data collection. Data were abstracted within 30 days from multi-chart and registration book for socio-demographic, baseline characteristics, follow up and outcome status by using BSC nurses as data collectors from nearby health centers. Two public health officers were used as supervisors.

### Data quality management

To ensure quality data collection, close supervision was carried out by the principal investigator and a supervisor during data abstraction. In addition, training was given by the principal investigator for two days before data collection period about the objectives of the study, variables on the study abstraction sheet, Stabilization center cards/multi-chart/ and how to abstract data for this study. The completed data abstraction form was checked for completeness of information by supervisors and principal investigator on daily basis.

### Data processing and analysis

Data was entered into STATA version 15 for analysis. Logistic regression was used to analyse treatment outcome with exposure variables. Variance inflation factor (VIF) was used to measure multicollinearity. Multinomial logistic regression was carried out to variables identify statistically significant variables at regression (p value <0.05).

### Ethical considerations

Ethical clearance was obtained from the Arba-Minch University Research and Ethics Publication Committee (RPC) to conduct the study. Further permission was obtained from zonal health department and medical director of the hospital. Confidentiality was maintained by making the data collectors aware of not to either expose or record any identification information found on the card and registration book.

## RESULT

### Socio-demographic characteristics of children admitted with SAM

A total of 453 medical records of children admitted with SAM to Jinka general hospital were reviewed.

Among the children admitted with SAM to the hospital were males compared female children attend. In terms of age proportion, the higher admissions fall in the range of 6 to 59 months.

Significant number of children with SAM were admitted from rural areas in comparison with the children from urban areas. Furthermore, the study shows that only small number of children fed on exclusive breast. This could be associated with increment in SAM admission to the hospital due unhygienic weaning diet preparation and lack of knowledge on diversification of weaning diet. This was further linked to lack of literacy among majority mothers which accounts 99 %. These findings align with underlying determinates outlined in the UNICEF Conceptual Framework on Maternal and Child Nutrition such as nutrient rich food to children with safe drinking which depends on household feed security. Adequate food preparation with hygienic conditions embedded into sanitation, adequacy of nutrition, maternal education affect the services to children at the household level (22).

The study also found that some dehydration was the major clinical manifestation associated diarrhea among the admitted SAM children. Kwashiorkor was the major clinical SAM among the admitted children to the hospital. This highlights that lack of excusive breast feeding has the potential to expose children to initiation of nutritionally poor staple foods.

In summary, altered vital signs such as respiration rate, temperature, and pulse rate were the major vitals assessed in Muli charts of children admitted with SAM. However, all 453 medical records such as multi charts were incomplete which shows poor monitoring of children admitted with SAM to the hospital. This needs attention of the medical personals to closely monitor the SAM children for better outcomes of the treatment. Furthermore, majority of the admissions were new ones to the hospital.

**Fig 1:**
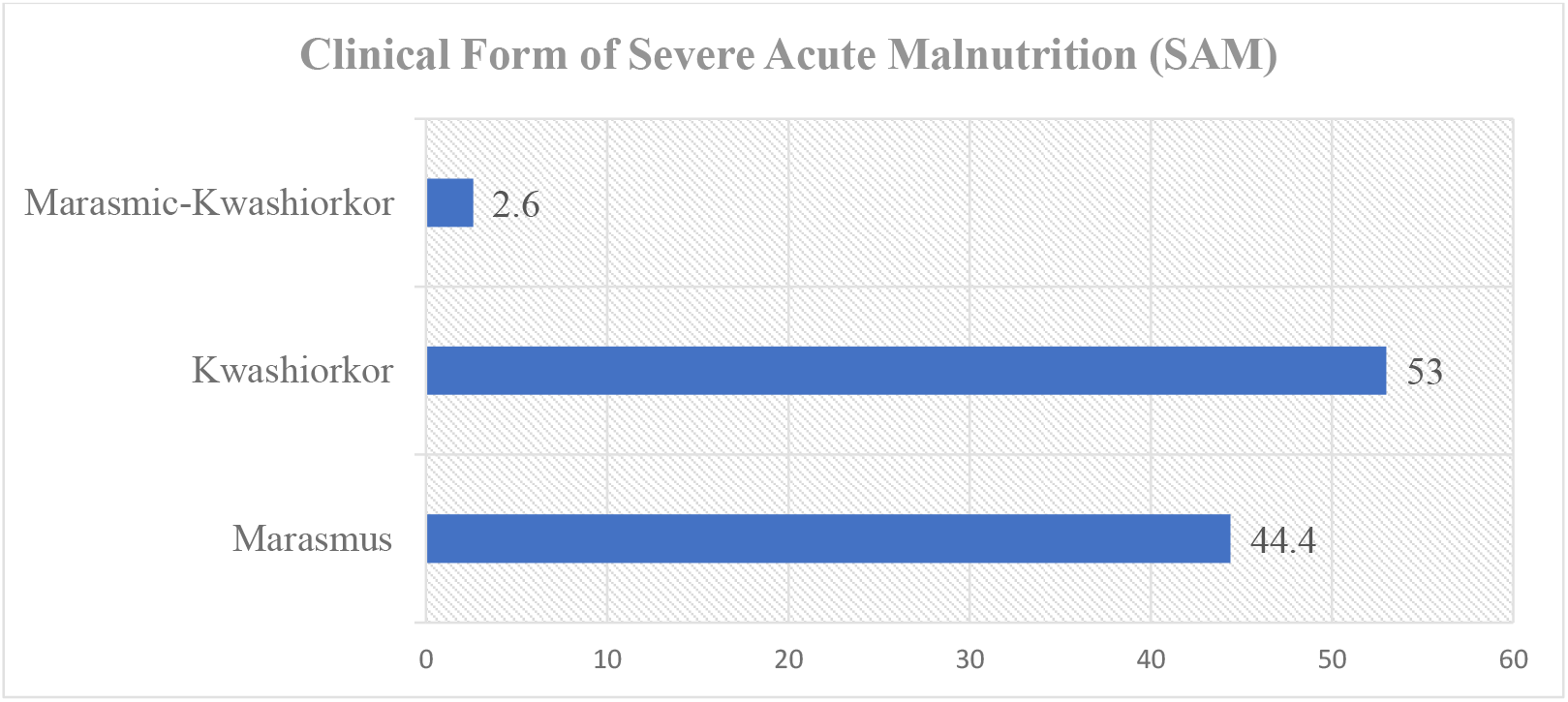
Clinical form of SAM of among children admitted Jinka General Hospital.

### Medical complication

Nutritional edema, shock, anemia, pneumonia, dermatosis, and consciousness were majorly recorded among the admitted SAM children at variables proportions. To provide medical care various laboratory investigations were conducted blood film, stool, urine analysis, haematocrit, blood glucose test, and HIV test where stool examination was the major laboratory investigation.

### Medication and Therapeutic feeding

Amoxicillin, Ampicillin, Gentamycin and Ceftriaxone were used at large scale which demonstrate that there was a high prevalence of medical complications among the children admitted with SAM. Likewise, F-75 and F-100 therapeutic milks were used at stabilization phased and rehabilitation phase to treat SAM.

The study also showed that blood transfusion and IV fluid administration medical orders were applied at low scale. This indicates that severe anemia prevalence among the children admitted with SAM and low medical complications that require IV fluid infusions despite 43.3% of cased managed with NGT for feeding during the stabilization phase. Among the admitted children a very few children 3% could not respond to the treatment during the medical in the hospital. This could be associated with underlying medical conditions that required further advanced medical care and intervention.

### Treatment outcomes

The finding of the study shows that majority of the admitted children with SAM were cured. This indicates that more than half of the admitted children cured from SAM (median, 1) and below of the admitted children treatment outcome was unsuccessful (median,0). However, it the cure rate was the below the SPHERE standard of acceptance of cure rate which prescribes >75%. Similarly, the death rate among the admitted children was at alarming rate of according to SPHERE standard of treatment outcomes. On the other hand, the default rate falls within the acceptable range of SPHERE standard whereas a few children with SAM were found be non-responders to the medical care. Similarly, only child was medically transferred to other medical facility for better medical intervention.

The highest standard deviation indicated that majority of the children treatment outcome was successful; however, 35% of the admitted children treatment outcomes were unsuccessful.

In summary, the successful treatment outcome (cured) was below the standard of SPHERE and death rate was at alarming range. These indicates that medical care of children admitted with SAM to the hospital should be improved through adhering with national treatment protocols. The poor medical care was also evidenced by incompleteness of multi chart of SAM children. Table one shows the descriptive statistics.

**Table one:** descriptive statistics.

| **Treatment outcome** | **observation** | **Mean** | **Percentage** | **Std. Dev.** | **Median** | **Mode** |
| --- | --- | --- | --- | --- | --- | --- |
| Cured | 453 | .6512141 | 65 | .477113 | 1 | 1 |
| Dead | 453 | .1567329 | 15.7 | .3639507 | 0 | 0 |
| Defaulted | 453 | .0441501 | 4.4 | .2056556 | 0 | 0 |
| Medical transferred | 453 | .0110375 | 1.1 | .2056556 | 0 | 0 |
| Non responded | 453 | .0309051 | 3.1 | .1732519 | 0 | 0 |

**Table two:** The comparison of the treatment outcome indicators (SPHERE standard)

| Indicators | Result | Outcome indicators for SAM |  |
| --- | --- | --- | --- |
|  |  | Acceptable | Alarming |
| Cure rate | 65 | >75% | <50 |
| Death rate | 15.7 | <10% | >15 |
| Defaulter rate | 4.4 | <15% | >25 |

### Factors associated with treatment outcomes of children admitted with SAM

The exit categories of children admitted with Sever Acute Malnutrition to hospital were analyzed successful (cured), unsuccessful (died) and unknown (defaulted, transferred out, non-responder). The STATA output showed that medications, medical complications, admission type, clinical form of SAM and socio demographic characteristics were the determining factors of treatment outcomes.

#### Ordered logistic regression (cured treatment outcome)

The result showed that successful treatment outcome of children with SAM was associated with variables such as therapeutic milk F-100, Gentamycin, sex of the child (male), rural residence, new admission, bilateral Pitting Oedema +++(Kwashiorkor), some dehydration. Children with new admission had a successful treatment outcome (cured). These variables significantly increase the likelihood of successful treatment outcome of children admitted with SAM to the hospital. Bilateral Pitting Oedema +++(Kwashiorkor), some dehydration and therapeutic milk F-100 are the key predictors of recovery of SAM cases.

**Table one:** Shows predictors of recovery among children admitted with SAM at P-value <0.05

| Variables | Coef. | Std. Err. | t | P>\|t\| | [95% Conf. Interval] |
| --- | --- | --- | --- | --- | --- |
| F 100 | .9767442 | .0086304 | 113.17 | 0.000 | .9597825, .9937058 |
| Gentamycin | .0232558 | .0105293 | 2.21 | 0.028 | .0025623, .0439493 |
| Sex -Male | .6870748 | .0454543 | 15.12 | 0.000 | .5977425, .7764072 |
| Rural residence | .3129252 | .0666249 | 4.70 | 0.000 | .1819859, .4438645 |
| New admission | .2669903 | .1155516 | 2.31 | 0.021 | .0398971, .4940835 |
| Bilateral Pitting Oedema<br>+++ (Kwashiorkor) | .7330097 | .0525032 | 13.96 | 0.000 | .6298253 .8361941 |
| Some dehydration | .814433 | .035875 | 22.70 | 0.000 | .7439279 .8849381 |

**Table two:** Predictors of death among children admitted with SAM.

| Variables | Coef. | Std. Err. | t | P>\|t\| | [95% Conf. Interval] |
| --- | --- | --- | --- | --- | --- |
| Rehydration Solution for Malnutrition (ResoMal) | .7692308 | .0548209 | 14.03 | 0.000 | .6614895, .8769721 |
| Educational status of mother | .2857143 | .0488891 | 5.84 | 0.000 | .1896315, .3817971 |
| Exclusive breast feeding | .7142857 | .0338985 | 21.07 | 0.000 | .6476643 .7809072 |
| Returned defaulter | .9681818 | .1006547 | 9.62 | 0.000 | .7703689 1.165995 |
| Severe Wasting with Bilateral Pitting Oedema (Marasmus-Kwashiorkor) | .957672 | .0388872 | 24.63 | 0.000 | .8812484 1.034095 |
| Infants 0-6 Months of Age | .5 | .0433526 | 11.53 | 0.000 | .4147992 .5852008 |
| Unconscious | .952381 | .0285216 | 33.39 | 0.000 | .8963276 1.008434 |

#### Ordered logistic regression (death treatment outcome)

Rehydration Solution for Malnutrition (ResoMal), educational status of mother, exclusive breast feeding, returned defaulter, severe wasting with bilateral Pitting oedema (Marasmus-Kwashiorkor), Infants 0-6 Months of age, and Unconscious were the substantial predictors of death children admitted with SAM.

During the management of children admitted with SAM need adequate attention to monitor and manage Rehydration Solution for Malnutrition (ResoMal). Moreover, targeted intervention based on these predictors is imperative.

## DISCUSSIONS

This study was conducted on 453 who were admitted with severe acute malnutrition to Jinka General Hospital (JGH) from to assess treatment outcome and associated factors. Other similar studies conducted in Ethiopia showed that the majority (92%) of the admitted children were from 6-59 months of age [2,3,4,5,14,15].

Majority (53%) of the children included in this study had bilateral pitting oedema (Kwashiorkor) in comparison with other similar studies conducted in Ethiopia [14, 18].

In this study, the death rate was 15.7% among the children admitted with SAM to Jinka. This result was below the minimum standard of SAM performance indicators set by SPHERE project of less than 15% [1] and lower than other studies findings at Asian and African countries [11, 12, 13]. However, the result was higher than study conducted at Yirgalem hospital which was 3.9%. Possibly, this may be associated with medical complications and poor quality of care evidenced by incomplete multi-chart, prescription of drugs out of national SAM management protocol. Based on this study, the death rate was worse in relation to five studies conducted in Ethiopia [17, 18, 19, 20, and 21]. The finding of this study indifferent with the studies conducted at Jimma,Yirgalem and Woldia Ethiopia [14, 15, and 20] which identified hypoglycaemia, dehydration, lower respiratory infection and therapeutic milk. Thes studies did not specify classification of dehydration and type of therapeutic milk. However, in this study some dehydration and therapeutic milk F-100 identified as associated factors.

Children with age less than 6 months were to die earlier than children with age 6 and above months. This finding matches with report of other studies [5, 12, 14, and 17]. Children with hypothermia were more likely to die from severe malnutrition than others without hypothermia like other studies [3, 4, 6 and 15]. But, in this study the death was associated with Rehydration Solution for Malnutrition (ResoMal), educational status of mother, exclusive breast feeding, returned defaulter, severe wasting with bilateral pitting oedema (Marasmus-Kwashiorkor, infants 0-6 months of age, unconscious which was dissimilar with study conducted in Dilla that associated death with the medical complication hypothermia [18]. In this study recovery of children was associated with some dehydration This result was similar with study conducted at Mekele [3,14,]. This could be associated with targeted intervention of dehydration management was key factors in improving the outcome of severely malnourished children admitted to hospital [19].

The study also highlighted that likelihood of death among the children admitted with SAM Rehydration Solution for Malnutrition (ResoMal). This was in contrary to a study conducted in Uganda [13] where intravenous fluid infusion were significant predictors of mortality [3, 5, 13, and 18].

Moreover, studies conducted in Ethiopia and other African countries did not show exclusive breast-feeding history, mothers’ education status, returned after defaulter, unconscious and Severe Wasting with Bilateral Pitting Oedema (Marasmus-Kwashiorkor as the predictors of death among children admitted with SAM. Among the predictors mother’s education and exclusive breast feeding supports UNICEF’s conceptual framework of malnutrition [22].

Similarly, the study identified that gentamycin, male, rural residence, new admission, bilateral pitting oedema +++(Kwashiorkor) as the predictors of cure among the children admitted with SAM. Why the being male and residence in rural associated with better treatment outcome needs further investigation.

### Conclusion and Policy Implications

Management of children admitted with severe acute malnutrition to the hospital was below the treatment outcome indicators of both SPHERE standard and national treatment guideline standards.

This could be associated with low maternal education status and low breast-feeding practice that contributed to poor services to children at household level. The poor literacy level among the mothers also resulted in underlying determinates of children malnutrition where appropriate dietary practices, age-appropriate feeding pattern and adequate nutrition to the children are suggestive.

Children admitted with severe acute malnutrition medical complications needs targeted interventions for better treatment outcomes. This could include adequate monitoring and administration of ResoMal, adherence with treatment guidelines and supply chain management of the stabilization Centre. All medical records of children admitted with SAM were found to be incomplete which shows poor surveillance of children. This could be possible reason why the death rate was at alarming rate in the hospital.

Associated factors like male and living rural area were unknown why associated with cure of children.

### RECOMMENDATIONS

❖ Appropriate surveillance of children admitted with severe acute malnutrition is imperative to monitor the progress of treatment.
❖ Administration of Rehydration Solution for Malnutrition should be according to prescribed guideline to prevent fluid overload.
❖ Empowerment of women and girls is mandatory to improve their literacy to prevent underlying determinants of children malnutrition
❖ Demonstration of nutritionally rich weaning or complementary food preparation from locally available food sources improves provision of adequate nutrition and age preparation of age food.
❖ Provide health education to caretakers of children admitted with severe acute malnutrition is imperative to prevent readmission and household level causes of malnutrition.
❖ Technical support should be given to medical staffs providing care for children admitted with severe acute malnutrition according to National Guidelines for the management of acute malnutrition in Ethiopia.

## Data Availability

The data will be provided up on request of the author

## Declarations

‘**Clinical trial number: not applicable.’**

## Ethics approval and consent to participate

For this study ethical approval was granted by Arbaminch University. A consent of the Jinka hospital authorities was obtained from the Jinak general hospital authorities to access the medical records.

## Consent for publication

Not applicable

## Availability of data and material

Data are available from the author, Turufat Tukura Gemechu upon reasonable request

## Author’ contributions

Turufat designed the concept of study. Make statistical analysis, monitored data collectors and drafted the manuscript.

## Acknowledgements

The author the Jinak general hospital Chief Executive Officer (CEO) for their permission to access the medical records.

## Authors’ information (optional)

The article is an original and has not been submitted for publication elsewhere.

## REFERENCES

1. .EMOH(2016) Guidelinesfor the Managementof AcuteMalnutrition. Federal DemocraticRepublic of Ethiopia.

2. 2(UNICEF), U.N.C.s.F., Management of SevereAcute Malnutritionin children: Workingtowards results at scale. UNICEF ProgrammeGuidance Document, 2015: p. 124.

3. Melaku Gebremichael1, A.M.B., Mache Tsadik2, Treatment Outcomes and Associated RiskFactors of Severely Malnourished under FiveChildren Admitted to Therapeutic FeedingCenters of Mekelle City, Northern Ethiopia. Open Access Library Journal, 2014: p. 9.

4. Bekele3, T.G.M.K.B.T.H., Survival status and predictors of mortality among severely acute malnourished children under 5 years of age admitted to stabilization centers in Gedeo Zone: a retrospective cohort study. Therapeutics and Clinical Risk Management, 2017: p. 10.

5. Wegen Shiferaw(MD)1, B.T.M., Kolsteren Patrick(PhD), Tefera Belachew (MD), Treatment Outcome and Predictors of SevereAcute Malnutrition using the WHO Guideline at aReferral Hospital in Southern. ResearchGate 2015: p. 19.

6. Survey, E.D.a., Key Indicators Report. FEDERAL DEMOCRATIC REPUBLIC OF ETHIOPIA, 2016: p. 59.

7. series, L., Social and economic implications of malnutrition Lancet series 2013: p. 4.

8. project, C., The Cost of HUNGER in Ethiopia. 2012: p. 24.

9. A Joint Statement by the World Health Organization, t.W.F.P., the United NationsSystem Standing Committee on Nutrition and the United Nations Children’s Fund, COMMUNITY-BASED MANAGEMENT OFSEVERE ACUTE MALNUTRITION. 2007: p. 8.

10. Lindsey M Lenters1, K.W., Patrick Webb2, Tahmeed Ahmed3, Zulfiqar A Bhutta1,4*, Treatment of severe and moderate acutemalnutrition in low- and middle-income settings:a systematic review, meta-analysis and Delphiprocess. BMC Public Health, 2013. 13(Suppl 3):S23: p. 15.

11. Ashraf Uddin Ahmed1, T.U.A.MD. Sultan Uddin1, MD. Haider Alichowdhury2, Mohammed Hamidur Rahman3, Md Iqbal Hossain4, Outcome of Standardized Case Management of Under-5 Childrenwith Severe Acute Malnutrition in Three Hospitals of Dhaka City inBangladesh. BANGLADESH J CHILD HEALTH, 2013. 37 (1): p. 13.

12. Mahama Saaka, S.M.O., 2 Anthony Amponsem,1,2Juventus B. Ziem,1,2 Alhassan Abdul-Mumin,1,2 Prosper Akanbong,1,2Ernestina Yirkyio,2 Eliasu Yakubu,2 and Sean Ervin3, Treatment Outcome of Severe Acute Malnutrition Cases atthe Tamale Teaching Hospital. Nutrition and Metabolism, 2015.

13. Richard Nyeko*, V.C., Boniface Otto Ssegujja and Grace Flona Ayot, Treatment outcome among children underfiveyears hospitalized with severe acutemalnutrition in St. Mary’s hospital Lacor,Northern Uganda. BMC Nutrition, 2016. DOI 10.1186/s40795-016-0058-6: p. 7.

14. Habtemu Jarso*, A.W.a.F.A., Survival status and predictors of mortalityin severely malnourished children admittedto Jimma University Specialized Hospitalfrom 2010 to 2012, Jimma, Ethiopia: aretrospective longitudinal study. BMC Pediatrics, 2015. DOI 10.1186/s12887-015-0398-4: p. 13.

15. Alemneh Kabeta* and Gezahegn Bekele, A.K., Factors Associated with Treatment Outcomes of Under-five Children with SevereAcute Malnutrition Admitted to Therapeutic Feeding Unit of Yirgalem Hospital. Clinics in Mother and Child Health, 2017. DOI: 10.4172/2090-7214.1000261: p. 5.

16. Melkamu Merid Mengesha1 * N.D., Balewgizie SileshiTegegne1 and Yadeta Dessie1, Treatment outcome and factors affecting time torecovery in children with severe acute malnutritiontreated at outpatient therapeutic care program. Global Health Action, 2016: p. 10.

17. Efrem Teferi1, Meskele Lera1, Sahle Sita1, Zerihun Bogale1, Daniel Gemechu Datiko1, Mohammed Ahmed Yassin3, Treatment outcome of children with severe acutemalnutrition admitted to therapeutic feeding centers inSouthern Region of Ethiopia. Ethiop. J. Health Dev, 2010. 24(3): p. 234–238.

18. TADELE GIRUM ADAL, M.K., BEFIKADU Tariku, Incidence and predictors of mortality among Severe Acute Malnorished Under Five Children Admitted to Dilla University Referal Hospital:-A Retrospective Longtitudinal Study. Journal of Biology, Agriculture and Healthcare, 2016. 6(13).

19. Anna Berti a, b. c *, Enrico Rino Bregania c d, Fabio Manentia c, Cristina Pizzi, Outcome of severely malnourished children treatedaccording to UNICEF 2004 guidelines: a one-yearexperience in a zone hospital in rural Ethiopia. Transactions of the Royal Society of Tropical Medicine and Hygiene, 2008. 102.

20. Tefera Chane*1, L.O., Gudina Egata Atomesa2 and Eskezyiaw Agedew3, Treatment Outcome and Associated Factors among Under-Five Childrenwith Severe Acute Malnutrition Admitted to Therapeutic Feeding Unit inWoldia Hospital, North Ethiopia. Journal of Nutrition & Food Sciences, 2014. DOI: 10.4172/2155-9600.1000329: p. 6.

21. Oumer, A., Survival Status and Predictors of Mortality among Children Aged 0–59 Months Admitted with Severe Acute Malnutrition in Dilchora Referral Hospital, Dire Dawa from September 2011 to August 2015. 2016: p. 56.

22. Unicef (2021). Conceptual Framework on Maternal and Child Nutrition

